# Estimates of the Value of Life Lost from COVID-19 in Ohio

**DOI:** 10.1101/2020.10.27.20220921

**Authors:** Peter J. Mallow

## Abstract

The economic burden of mortality due to the novel coronavirus (COVID-19) was estimated for the State of Ohio. Data from the Ohio Department of Public Health and Social Security Administration was used to estimate the years of potential life lost, 56,518, and economic value of those lost lives, $13.60 billion. These estimates may be used to assess the risk-trade off of COVID-19 mitigation strategies in Ohio.

## Introduction

We have long recognized that the burden of a disease [i.e. the novel coronavirus (COVID-19)] extends beyond the number of individuals infected or who die. [1] Thus, we need to understand the economic value of those premature deaths as a society to make policy decisions about different mitigating strategies. [1,2] It has been estimated that mortality from COVID-19 has cut short approximately 2.5 million years of potential life in the United States. [3] However, the economic value of these lost life years was not captured, nor were local estimates of the years of potential life lost (YPLL) provided to aid state level policy makers, who have taken the lead in responding to COVID-19. [4]

COVID-19 has contributed to 5,078 Ohioans deaths as of October 20, 2020. [5] Though, the full burden caused by COVID-19 mortality is not captured solely by the lives lost. Each death represents some number of YPLL had the individual not contracted COVID-19. Further, the statistical value of those lives lost (VSL) is not captured in mortality data alone. The estimated YPLLs and associated economic value for Ohio were estimated using data from the Ohio Department of Health and United States Social Security Administration. [5,6] The results suggest Ohio suffered 56,518 YPLLs and an associated lost VSL of $13.60 billion dollars [Figure 1].

**Figure 1a.**
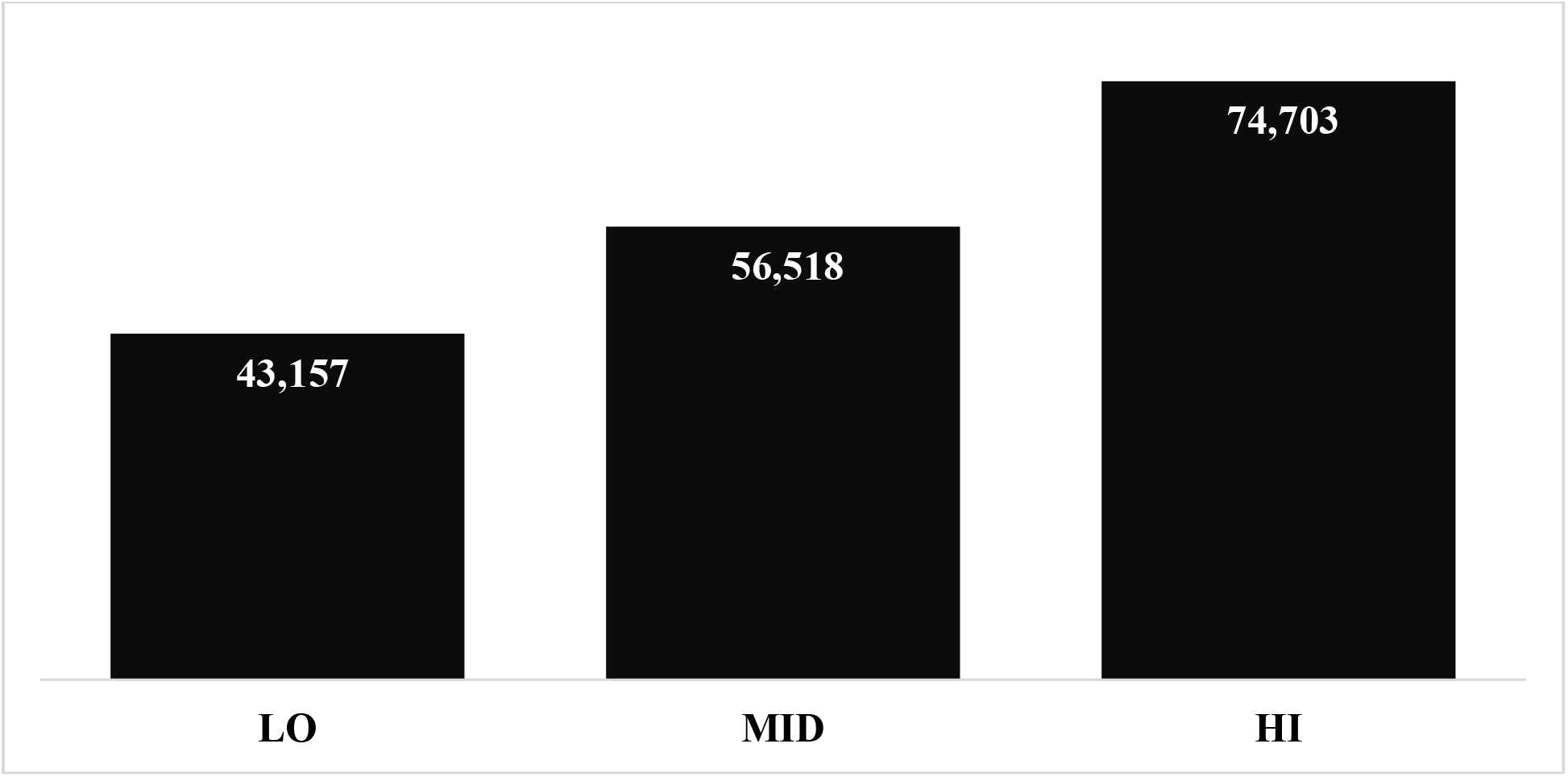
Years of Potential Life Lost (YPLL)

**Figure 1b.**
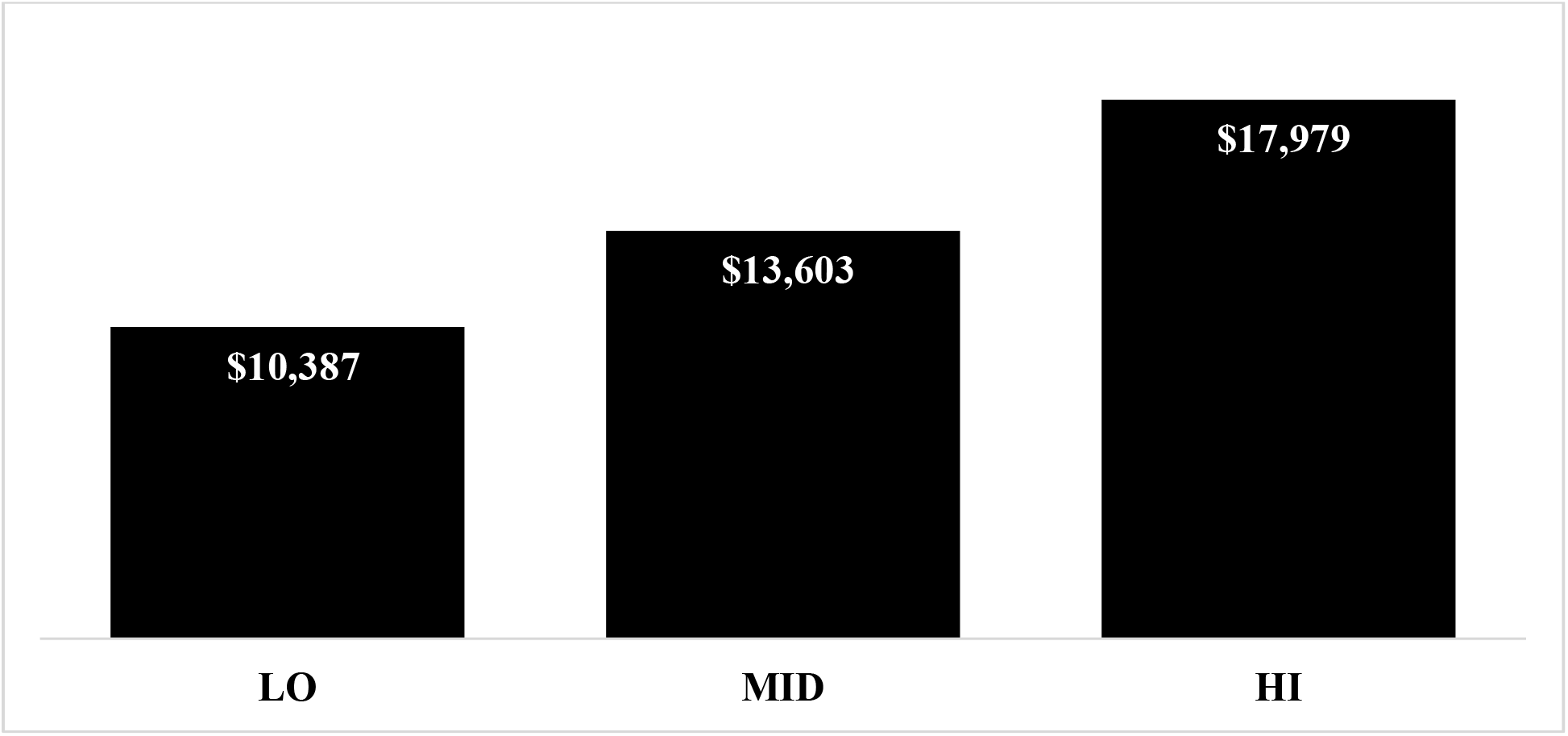
Value of Statistical Life (VSL) Lost, in billions.

### Data and Methods

Ohio COVID-19 mortality was downloaded on October 20, 2020 from Ohio Department of Public Health’s (ODPH) website. [5] The data included all known cases of COVID-19 including hospitalizations and deaths by county of residence, age range, and sex of the individual. The data is updated daily and changes often occur to past data based upon new information received by the ODPH. The age ranges were: 0-19, 20-29, 30-39, 40-49, 50-59, 60-69, 70-79, and 80+. The mid-point of the range was used to impute the age of death and the low- and high-end of the range was used to inform the sensitivity analysis. For those 80+, the mid-point was assumed to be 87 and the high-end to be 95. Those deaths with missing data, five individuals, were removed from the analysis.

Actuarial life tables were downloaded from the Social Security Administration (SSA) to determine the remaining years of expected life by age. [6] The life tables provided estimates of remaining life expectancy by age and by gender. The most recent life tables were published in 2017. The SSA used data published in the volumes of Vital Statistics of the United States and tabulated by the National Center for Health Statistics (NCHS). [7]

The YPLLs were calculated by subtracting the expected life expectancy from the imputed age of death by individual. The value of a life year lost was $247,676, and it was obtained from previous published literature. [8] The value of a life year lost, referred to as value of statistical life year (VSLY), is an estimate of the society’s willingness-to-pay for one year of life, and is used often by state and federal policymakers to quantify the burden of disease or policy. [1] The VSLY was multiplied by the YPLL to determine the economic burden associated with the lost value of life (VSL) for each COVID-19 death. The YPLL and VSL results were summed to generate aggregate results for the state, gender, and age (less than 60 & 60+).

## Results

The 5,078 deaths in Ohio resulted in an expected YPLL of 56,518 (range: 43,157 to 74,703). The corresponding VSL was $13.60 billion (range: $10.39 to $17.98 billion) [Figure 1]. Based on the age of the COVID-19 deaths, the average loss of potential life years was 11.2 per person (range: 8.6 to 14.8 years). The breakdown by gender was 26,327 and 30,192 YPLLs for females (2,478 deaths) and males (2,600 deaths), respectively. The economic loss by gender was $6.34 (female) and 7.27 (male) billion [Figure 2]. Estimates by age indicated that 13,123 of the 56,518 YPLLs and $3.16 billion VSL resulted from COVID-19 deaths under 60 [Figure 3]. For those deaths under the age of 60, the average loss of potential life years was 31.0-person years (range: 27.6 to 35.3). Whereas, the years of potential life lost in deaths aged 60 or greater was 9.4-person years (range: 6.8 to 13.0).

**Figure 2a.**
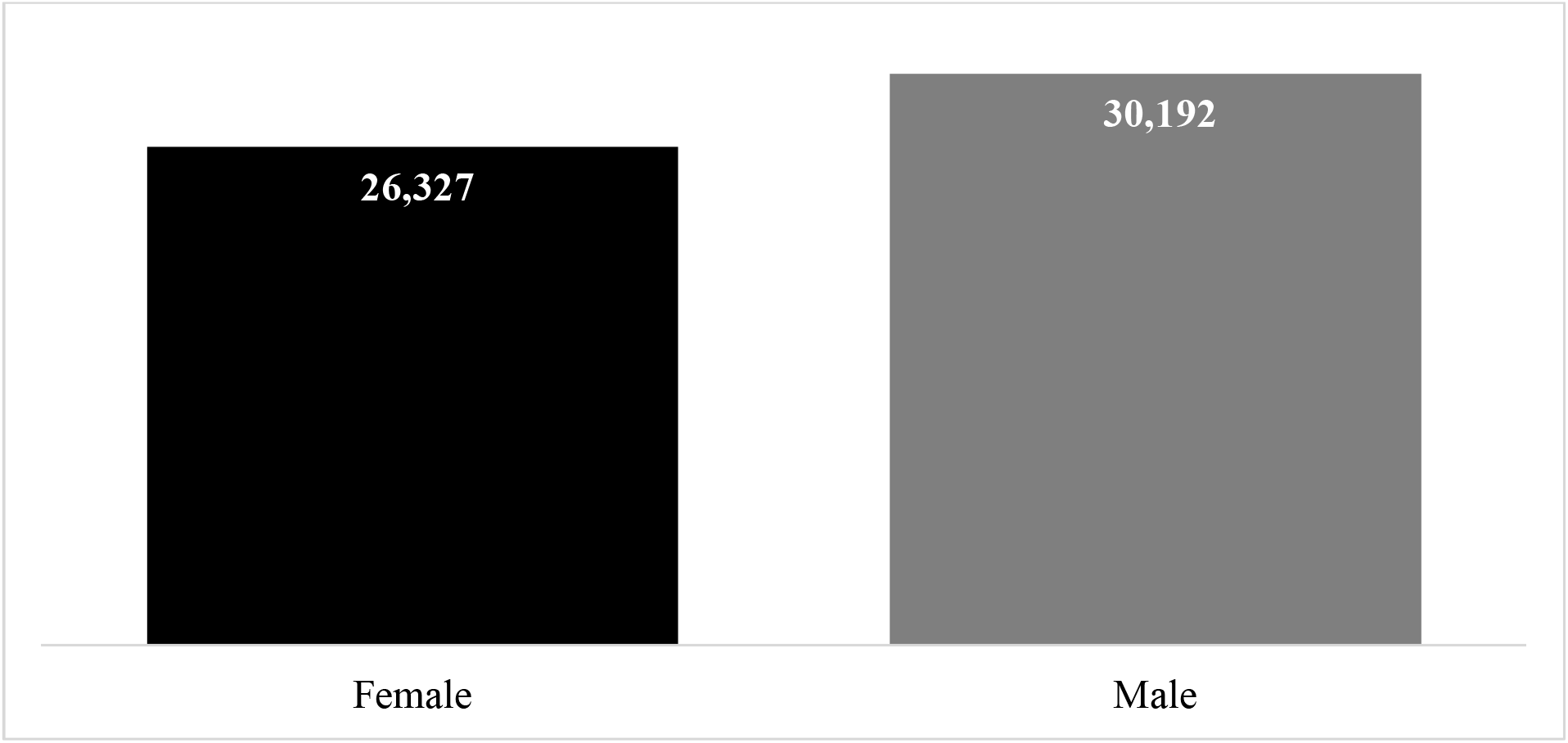
Years of Potential Life Lost (YPLL) by Gender.

**Figure 2b.**
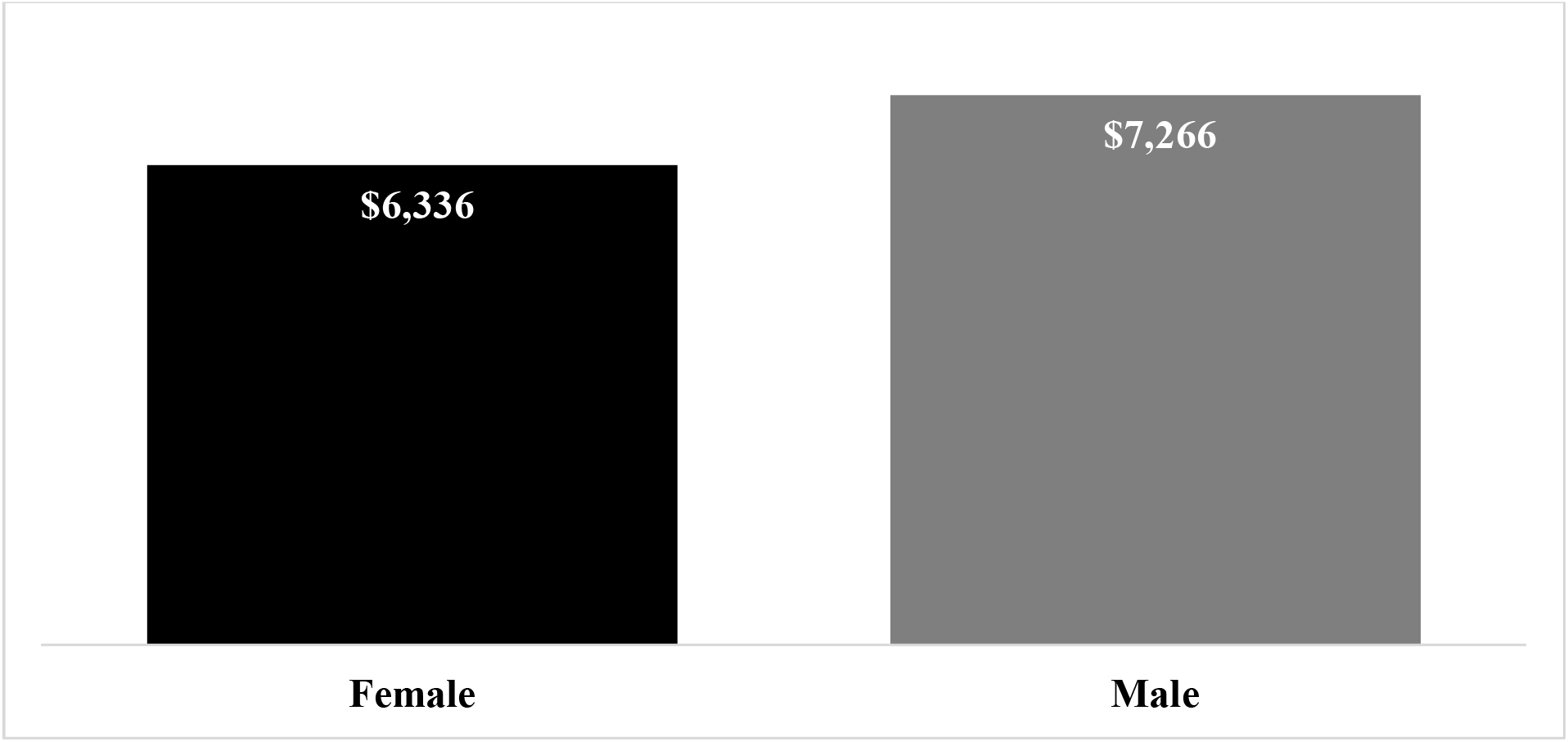
Value of Statistical Life (VSL) Lost by Gender, in billions.

**Figure 3.**
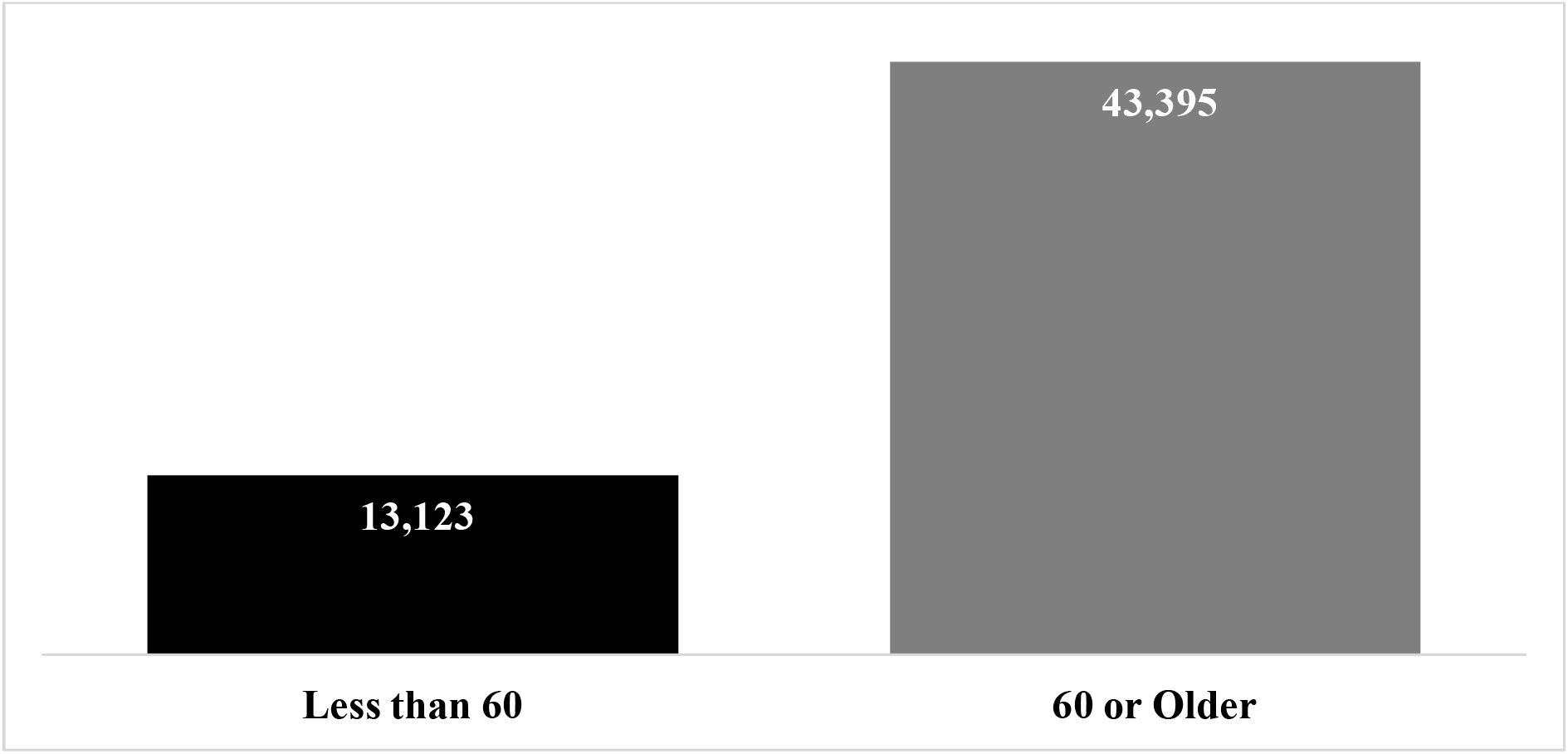
Years of Potential Life Lost (YPLL) by Age.

**Figure 3b.**
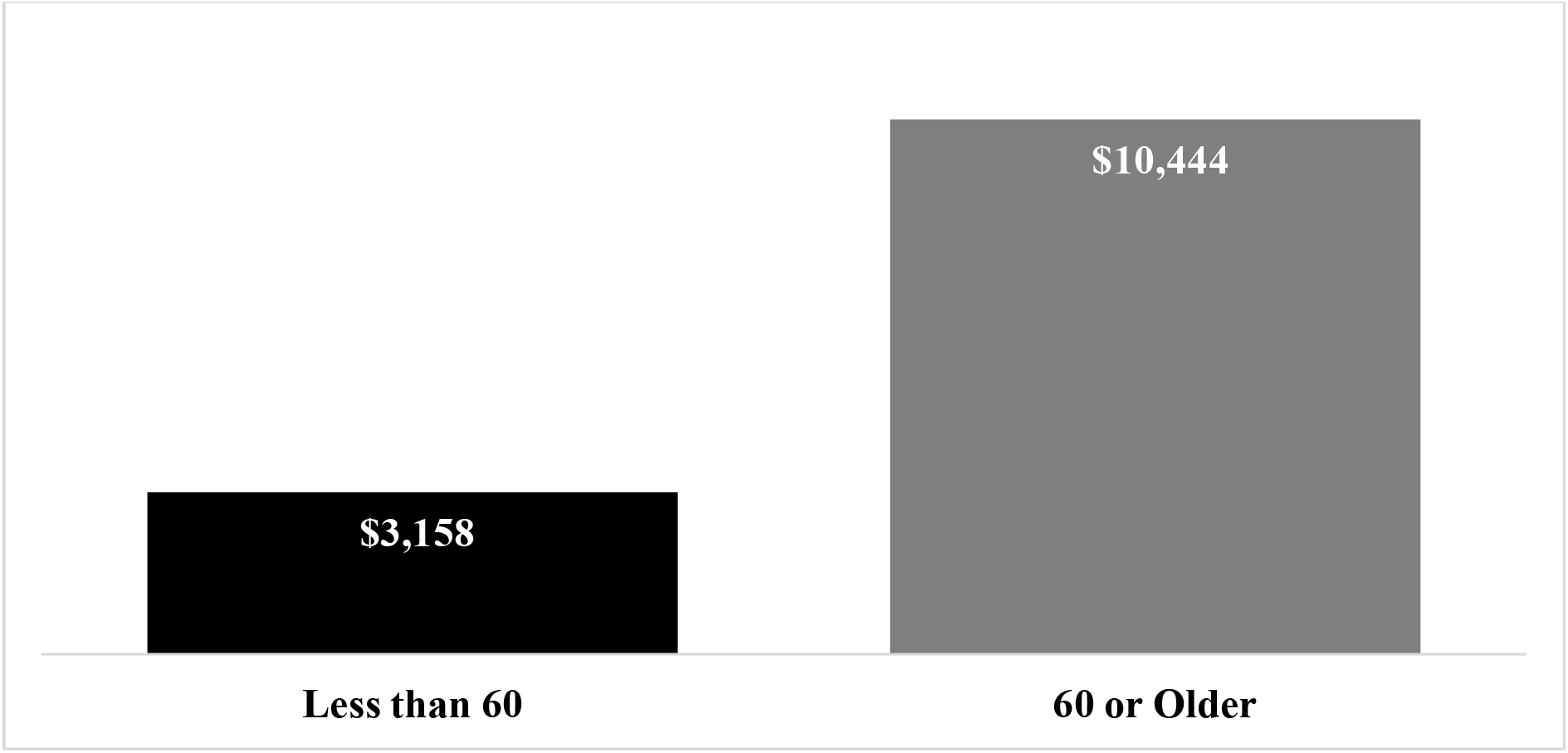
Value of Statistical Life (VSL) Lost by Age, in billions.

## Discussion

Building upon the work conducted for the nation, this study estimated the YPLL and VSL due to COVID-19 in Ohio. [3] Assuming a consistent value of a year of lost life, the economic burden of premature deaths was estimated at $13.60 billion as of October 20, 2020. This value and the estimated YPLLs are rapidly changing with the current spread COVID-19 in the community. Thus, the results likely underestimate the YPLLs and VSL of the loss of life from COVID-19 in Ohio.

Separating the results by gender indicated that the age distribution and corresponding YPLLs and VSL were similar (46.6% female; 53.4% male). An examination by age indicated the disproportionate effect of those under COVID-19 deaths under 60 has on the expected overall results. Of the 5,078 deaths, 424 (8.3%) were under 60. However, these deaths represented 23.2% of the total YPLLs and VSL. To date COVID-19 mortalities under 60 have been relatively few, the much greater expected loss of potential life years, 21.6-person years, contributed to the higher YPLL and VSL.

These results have implications for state level policies addressing the COVID-19 pandemic. The estimates identified in this study for Ohio may provide a common and consistent framework to estimate the costs and benefits of differing COVID-19 mitigation strategies, such as social distancing, vaccines, and novel treatments. [1]

The method of estimating the VSL for a particular health condition is not without controversy. [8, 9] The methodology used assumes a standard value of life across all individuals. As a result, allocation and equity issues associated with the assessment of differing policies may vary by age, income, and race. This approach should be one of several when assessing strategies to mitigate the deleterious effects of COVID-19. Further, the ODPH does not release the specific age of the individuals who have died from COVID-19, which resulted in imputations to estimate the YPLLs and VSL. This limitation was mitigated by sensitivity analysis examining the low- and high-end of the age ranges. Finally, there is no generally accepted appropriate VSLY. The use $247,676 in this study was transparent and the method allows for the ease of application of differing amounts.

The expected loss of life-years from premature deaths of COVID-19 in Ohio was substantial. Each death represents a number of life-years lost and the enjoyment of those years with family and friends. Further, there is a substantial economic loss to all Ohioans from each death. Estimating the economic burden of COVID-19 may provide a consistent means for state level policy makers to assess interventions control and mitigate the effects of the COVID-19 pandemic.

## Data Availability

The data will be made available upon reasonable request.

## Institutional Review Board (IRB)

This research was deemed exempt from IRB review by Xavier University.

